# Long-term exposure to air pollution impacts activity in brain regions involved in inhibitory control in 10 to 13- year old children

**DOI:** 10.1101/2025.11.26.25341043

**Authors:** Mikołaj Compa, Yarema Mysak, Clemens Baumbach, Paulina Lewandowska, Aleksandra Domagalik, Bartosz Kossowski, Katarzyna Kaczmarek-Majer, Anna Degórska, Krzysztof Skotak, Katarzyna Sitnik-Warchulska, Małgorzata Lipowska, Bernadetta Izydorczyk, James Grellier, Iana Markevych, Marcin Szwed

## Abstract

**Background:** The development of inhibitory control, a core component of cognitive control, can be influenced by environmental factors. We investigated whether exposure to particulate matter with diameter ≤ 10 μm (PM_10_) and nitrogen dioxide (NO_2_) across different life-time periods is related to the neural correlates of inhibitory control in 10-to 13-year-old children from southern Poland.

**Methods:** Task functional magnetic resonance imaging (task fMRI) measures brain activity while participants perform specific cognitive or behavioral tasks. We investigated inhibition using a Go/NoGo task during task fMRI and tested associations between neural correlates of inhibitory control and exposure to air pollution during prenatal, early-life, and current life periods. The study population comprised children from the NeuroSmog study with Attention-Deficit/Hyperactivity Disorder (ADHD, n=143) and their typically developing peers (n = 385).

**Results:** Higher current exposure to PM_10_ was significantly associated with reduced brain activation during response inhibition in key cognitive control networks, including the dorsolateral prefrontal cortex and anterior cingulate cortex. We did not observe significant interactions between our participants’ ADHD diagnosis and their exposure to air pollution.

**Conclusions:** Long-term exposure to air pollution was associated with impairments in brain function related to inhibition in both ADHD children and their typically developing peers. Our findings add novel, pertinent evidence to the growing body of research indicating that air pollution negatively impacts the development of executive function in children and suggests that the same mechanisms that underlie pollution’s effects on the brain may also lead to the increased incidence of ADHD.

## Introduction

Air pollution is the leading environmental risk to human health (Brauer et al., 2024). Exposure to air pollutants is related to increased rates of cardiovascular and pulmonary diseases such as stroke, elevated blood pressure, and chronic obstructive pulmonary disease (De Bont et al., 2022; Duan et al., 2020; Konduracka & Rostoff, 2022). Inhalation of air pollutants can also negatively impact the central nervous system (Morrel et al., 2025), an effect that may be explained by a dysregulated immune response. One of its mechanisms involves pollutants activating microglia, which triggers chronic neuroinflammation (R. Babadjouni et al., 2018; R. M. Babadjouni et al., 2017; Elder et al., 2004; Habre et al., 2018; Li et al., 2017; Wilson et al., 2010). Over time, this sustained inflammatory state can damage neurons and decrease overall brain network activity.

Exposure to air pollution has been linked with poorer performance in several cognitive tasks (Compa et al., 2023; Forns et al., 2016, 2017; Gignac et al., 2021, 2022; Guxens et al., 2014; Saenen et al., 2016; Sunyer et al., 2015, 2017). Chronic exposure to particulate matter (PM) and NO_2_ is associated with poorer sustained and selective attention (Saenen et al., 2016), slower cognitive development trajectories in working memory that persist for years (Forns et al., 2017; Sunyer et al., 2015), and more behavioral problems in schoolchildren (Forns et al., 2016). The effects may begin even before birth, as prenatal exposure to NO_2_ has been linked to delayed psychomotor development (Guxens et al., 2014). Short-term and exposure to nitrogen dioxide (NO_2_) has also been associated with attention deficits in schoolchildren, such as slower response times and more errors (Sunyer et al., 2017), and with less efficient executive attention in both typically developing (TD) children and children with Attention-Deficit/Hyperactivity Disorder (ADHD), a neurodevelopmental condition characterized by persistent patterns of inattention, hyperactivity, and impulsivity (Compa et al., 2023). In addition, multiple studies have found associations between exposure to air pollution and the incidence of ADHD (Abid et al., 2014; Kim et al., 2021; Markevych et al., 2018; Thygesen et al., 2020), further demonstrating the negative effects of exposure to air pollution on cognitive development.

Detrimental effects of exposure to air pollution have been observed not only in strictly behavioral studies, but also in neuroimaging studies. However, the effects on brain structure, while investigated in multiple studies, have turned out to be often inconsistent (see recent systematic review by Morrel et al. (2025). Effects on brain function on the other hand, offer a clearer and more consistent picture, suggesting that alterations in brain activity could be a better indicator of negative effects of exposure to air pollution.

Several studies indicate that air pollution disrupts the maturation of large-scale brain networks by interfering with normal integration and segregation processes (Cotter et al., 2023; Gawryluk et al., 2023; Pujol et al., 2016; Zundel et al., 2024b). Integration involves strengthening connections within networks, whereas segregation reflects the pruning of connections between networks—both essential for typical neurodevelopment (Compa et al., 2023; Forns et al., 2016, 2017; Gignac et al., 2021, 2022). Higher exposure to pollutants such as PM2.5 has also been linked to elevated connectivity between the default mode network (DMN) and task-positive systems, suggesting impaired differentiation of these networks and delayed maturation (Cotter et al., 2023; Pujol et al., 2016; Zundel et al., 2024a). Acute diesel exhaust exposure can also reduce within-DMN connectivity (Gawryluk et al., 2023), and long-term pollution exposure appears to blunt the normal age-related strengthening of this network (Zundel et al., 2024). Although some of these effects may reflect compensatory reorganization, the overall evidence indicates that air pollution adversely alters the development of multiple brain networks.

Neuroimaging offers several methods of studying of how air pollution impacts the brain. Another method that is particularly relevant yet still underused in this field is task-based functional magnetic resonance imaging (task fMRI). Task fMRI measures brain activity while participants perform specific cognitive or behavioral tasks and allows one to identify neural systems engaged is a particular mental process.

The present study aimed to fill this critical evidence gap by examining associations between exposure to air pollution and task-related brain activity related to inhibitory control. To achieve this aim, we employed a classic Go/NoGo task during functional Magnetic Resonance Imaging (fMRI) to measure brain activity related to inhibition, an important component of cognitive control and executive attention (Badre, 2025; Matsumoto & Tanaka, 2004). Similar to other, pre-registered studies on this cohort (Lewandowska et al., 2025) we chose three time-windows of long-term exposure to air pollution – prenatal, early-life (0 to 4 years), and current (long-term exposure at current home address) (at time of recruitment). Prenatal and early-life periods are critical for the development of the central nervous system; we expected that the effects of exposure to air pollution during these time windows could have the largest impact (Guxens et al., 2018; Rivas et al., 2019). Finally, we explored whether the impact of air pollution on brain activity during the Go/NoGo attentional task is dependent on ADHD status. By testing for such an interaction effect, we aimed to determine if the brains of individuals with ADHD, an at-risk population, are more sensitive to exposure to air pollution than the brains of their TD peers.

## Methods

### Study area and population

Participants of the NeuroSmog study (Markevych et al., 2021) were recruited in the years 2020–2022 among children aged 10–13 years living in 18 towns located in the southern part of Poland. Towns were specifically chosen for their diverse air pollution levels and population sizes. Children at risk of ADHD diagnosis were recruited using convenience sampling and were referred to the study by local psychologists, while likely TD children were recruited using random sampling. Children with intellectual disability, neurological or psychiatric disorders, or other serious medical conditions were excluded. We also excluded children with a gestational age less than 35 weeks or birthweight smaller than 2000 grams. To be included, children and their parents had to be fluent in the Polish language and to attend a school in one of the study towns for at least a year prior to recruitment. Each child was subjected to psychological evaluation, including a full ADHD diagnosis. All testing was done by trained clinical child psychologists. Details on child selection and evaluation procedures are described in Markevych et al. (2021).

Initially, we recruited 741 children (217 with ADHD diagnosis and 524 TD). To create the analytic sample, we removed data of children who did not complete MRI scanning, performed overly poorly during the scanning task, who were outliers in air pollution or fMRI data, or who lacked data on demographics or exposure to air pollution. This resulted in a sample comprising 545 participants for analyses on current long-term exposure to air pollution and a sample comprising 528 participants for analyses on pregnancy and early-life exposure to air pollution (Figure 1).

**Figure 1.**
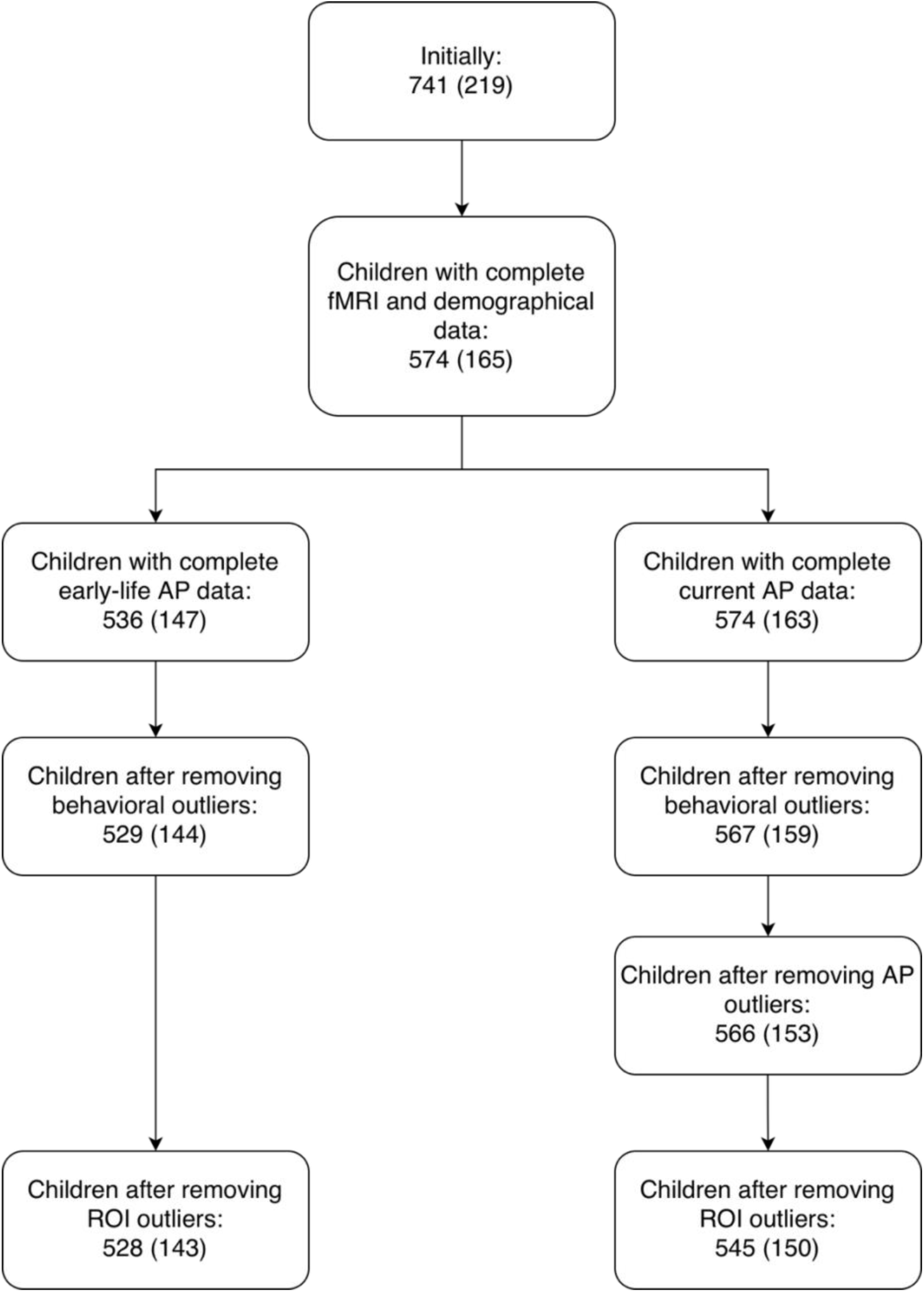
A flowchart showing the construction of the two analytic samples. Numbers in parentheses represent the number of children with ADHD diagnosis. Abbreviations: fMRI, functional Magnetic Resonance Imaging; AP, Air Pollution; ROI, region of interest.

The study was conducted according to the guidelines of the Declaration of Helsinki and was approved by the Ethics Committee of the Institute of Psychology, Jagiellonian University, Kraków, Poland (#KE_24042019A). Written informed consent was obtained from the legal guardians, and all children gave their written informed assent. The Clinical Trials Identifier is NCT04574414.

### Air pollution exposure assessment

Estimates of mean air pollution concentrations were derived from hybrid land use regression (LUR) models. The resulting raster maps of mean NO_2_ and mean PM_10_ concentrations in the study area had a spatial resolution of 125 m × 125 m and were available for each month from 2007 to 2012 and for each year from 2013 to 2018. Following the statistical approach outlined by de Hoogh et al., 2018, stepwise forward linear regression models were applied. These models were used to estimate air pollution levels based on ground measurements and influencing predictive factors from multiple sources. The main groups of predictor variables considered in the LUR models included: (i) traffic and residential emissions of air pollutants from dispersion models and (ii) land-use data from the CORINE land cover database (European Environment Agency, 2018) and the Polish Database of Topographic Objects (BDOT10k) (Sęp & Sikora, 2018). Significant predictor variables were included in the model if their addition resulted in an increased adjusted R^2^ relative to the previous step. The LUR models were validated using 5-fold cross-validation. The adjusted R^2^ values indicating the model fits for 2018 were 0.85 for NO_2_ and 0.64 for PM_10_. For the years spanning the early-life period (2007–2017), the average adjusted R^2^ was 0.82 (with range 0.64−0.92) for the NO_2_ models and 0.66 (with range 0.43−0.84) for the PM_10_ models.

Current long-term exposure to air pollution was defined as the annual mean air pollution concentration of 2018 at the address where the child was living at the time of recruitment. Prenatal exposure was calculated by averaging the monthly mean air pollution concentrations at the mother’s pregnancy address or addresses over the last two trimesters of the pregnancy as these are the periods most relevant for brain development. The start of pregnancy was determined by subtracting the reported gestational age from the child’s birthdate; in cases of missing gestational age, 40 weeks were assumed. Early-life exposure was calculated by averaging the monthly and/or annual mean air pollution concentrations over the child’s home addresses from birth to the child’s fourth birthday.

### MRI procedure

#### Mock scanner training

To ensure the best possible quality of MRI images, we employed a mock scanner protocol in our study (Suzuki et al., 2023). Children underwent a 15-minute mock scanner session to get prepared for a real MRI. They first acclimated to the loud noise and confined space. Using a motion sensor with visual feedback, they practiced lying still. They then completed a three-stage task training, progressing from practice with feedback to a final test requiring 80% accuracy without feedback. This process ensured that each child was comfortable and ready for the actual scan.

### Go/NoGo task

During fMRI, each child performed a Go/NoGo task. The task performed in the scanner was based on the Human Connectome Project’s Conditioned Approach Response Inhibition Task (CARIT) task (Somerville et al., 2018). Participants were asked to react to Go stimuli and to ignore NoGo stimuli. Each stimulus was displayed for 600 ms. The interstimulus interval was jittered and lasted between 1 and 4.5 seconds. The task included 68 (74%) Go and 24 (26%) NoGo stimuli. In total, two runs of the task were completed, each lasting 4 minutes and 19 seconds. The number of Go stimuli preceding a NoGo stimulus, known as prepotency, was set to 2, 3, and 4, and distributed randomly throughout the task. The Go/NoGo task generates four types of events: a reaction to a Go stimulus is called a hit; a non-reaction to a NoGo stimulus is a correct rejection; a reaction to a NoGo stimulus is a commission error; and a non-reaction to a Go stimulus is an omission error.

#### Image acquisition

All MRI data were acquired on the same Siemens Magnetom Skyra 3T scanner with a 64-channel head coil. Scans were performed at the Małopolska Centre of Biotechnology, Jagiellonian University in Krakow, Poland. Anatomical scans were acquired using the T1-weighted sequence to perform detailed alignment between the structural and functional scans. In this sequence, repetition time (TR) was 2500 ms, echo time (TE) was 2.9 ms, the flip angle was 8°, voxel size was 1 mm × 1 mm × 1 mm with 50% distance factor, and Field of View (FoV) was set to 256 mm. Slices were acquired in the interleaved sequence with the GRAPPA acceleration factor set to 2 (Feinberg et al., 2010; Moeller et al., 2010; Xu et al., 2013).

Two T2*-weighted sequences were used to acquire functional scans. In each sequence, TR was 800 ms, TE was 30 ms, and the flip angle was set to 52°. Voxel size dimensions were 2.4 mm × 2.4 mm × 2.4 mm, with FoV set to 210 mm. Sixty slices were collected in an interleaved fashion using the multiband acceleration factor of 6. Each of the two runs of the sequence lasted 4 minutes and 19 seconds. Both anatomical and functional sequences used anterior-posterior phase encoding direction. Both sequences were adapted from the ABCD study (Casey et al., 2018).

### Preprocessing of MRI data

Functional and structural data were preprocessed using fMRIPrep version 23.2.0 (Esteban et al., 2019) a Nipype-based tool (Gorgolewski et al., 2011). fMRIPrep’s preprocessing pipeline included skull-stripping, motion correction, slice-timing correction, co-registration to the participant’s anatomical image, and normalization to Montreal Neurological Institute (MNI) standard space. fMRI data was smoothed using a 7.2 mm kernel as implemented in the SPM12 toolbox (Wellcome Trust Centre for Neuroimaging, 2014). A comprehensive description of the pipeline can be found in Section 1 of the Supplementary materials.

### Statistical analysis

#### Identification of brain activity related to successful inhibition

For each participant and voxel, the observed blood-oxygenation-level-dependent (BOLD) signal time-series was modeled as a function of several regressors using a General Linear Model (GLM), as implemented in the SPM12 software package. Event-specific regressors for hits, correct rejections, commission errors, and omission errors were created by convolving the event-specific SPM12 canonical hemodynamic response functions (HRF). In addition to these event-specific regressors, the model included six motion parameters and the signal from the cerebrospinal fluid (CSF) as nuisance regressors.

To identify brain activity related to successful inhibition, we computed, for each participant and voxel, t-contrast images of correct rejections versus hits as the difference between their respective coefficient estimates from the GLM. Positive contrast values represent the magnitude of the successful inhibition effect on brain activity.

In a second step, these individual-level contrast images were used in a group-level one-sample t-test. This test was performed at each voxel to determine if the group mean effect was significantly different from zero. To correct for multiple comparisons, we applied a standard SPM12 cluster-level family-wise error (FWE) correction based on Random Field Theory (RFT). This procedure involved two steps: first, we formed voxel clusters by grouping together voxels that shared a face and had uncorrected t-test p < 0.001. Second, RFT was used to produce cluster-level FWE-corrected p-values based on cluster extent. Only clusters with p < 10^-6^ were considered as showing significant activation related to inhibition and kept as candidates for further analysis.

#### ROI selection

Studies that used the Go/NoGo task report several brain regions involved in inhibition (Aron et al., 2014; Botvinick et al., 2001; Cai et al., 2014; Corbetta & Shulman, 2002; Criaud et al., 2020; MacDonald, 2000; Mostofsky & Simmonds, 2008; Simmonds et al., 2008). Hence, from our previously identified clusters (see paragraph above) we selected clusters spatially overlapping with brain regions reported in this literature and use those overlapping regions of interest (ROIs) in this study. This procedure resulted in 11 ROIs from the following brain regions: bilateral supramarginal gyrus, bilateral anterior cingulate cortex (ACC), bilateral dorsolateral prefrontal cortex (DLPFC), right hippocampus, right superior parietal lobule, bilateral anterior insula, and right pre-supplementary motor cortex (Pre-SMA). Each ROI was centered on its cluster’s peak activation voxel. Table 2 lists the ROIs and their centers’ coordinates.

#### Analysis of exposure to air pollution and ROI activation

To derive a single value that would represent a participant’s inhibition-related activation in a given ROI, we placed virtual 6 mm spheres around the ROI’s center and computed the average correct-rejection-versus-hit t-contrast over all the voxels within the sphere. This resulted in one value per participant and ROI. These values were then used as dependent variables in separate, ROI-specific linear regressions. The primary predictor of interest in those regressions was air pollution exposure for each pollutant (NO_2_ and PM_10_) and for each time window. Covariates were selected as described in our previous research (Compa et al., 2023; Lewandowska et al., 2025; Szwed et al., 2025) and included sex, exact age in years, ADHD status, and town size (“large” if ≥ 90,000 inhabitants, otherwise “small”, which is in line with the NeuroSmog study’s design (Markevych et al., 2022)). We also ran a sensitivity analysis with an additional adjustment for socioeconomical status (SES) approximated by the parents’ minimum level of education (“low” if primary or vocational education, “medium” if high school and additional training, “high” if Bachelor’s degree or higher). To assess the robustness of these models to the specific definition of the individual ROI, we repeated the full analysis using data extracted with two alternative methods: using larger spheres (8 mm and 10 mm) and using average t-contrasts computed over voxels from the entire anatomical brain region containing the ROI’s center, e.g., the ACC.

Outliers were removed from the dataset based on visual inspection of histograms and scatterplots generated for each continuous variable. Generalized additive models were fitted and plotted to visually check for possible nonlinear relationships between ROI variables and exposures to air pollution while adjusting for sex, age, ADHD status, and town size (Hastie, 2017). Residual analysis was done using R’s DHARMa package (Hartig, 2017). Finally, we also ran models that included a two-way interaction for ADHD status and exposure to air pollution. To correct for multiple comparisons, we controlled the False Discovery Rate (FDR) at level α = 0.05 using the Benjamini-Hochberg procedure.

## Results

### Descriptive characteristics of the analytic sample

The study participants in the analytic sample had a mean age of 11.5 years, 228 of them were female, and 150 were diagnosed with ADHD (Table 1). Regarding the educational level, 42% were from families with a high minimum level of parental education. The mean concentrations of current, prenatal, and early-life PM_10_ were 36.5 μg/m^3^, 45.4 μg/m^3^, and 45.1 μg/m^3^, respectively (Table 2). The mean concentrations of current, prenatal, and early-life NO_2_ were 19.2 μg/m^3^, 23.8 μg/m^3^, and 24.47 μg/m^3^, respectively.

**Table 1.**
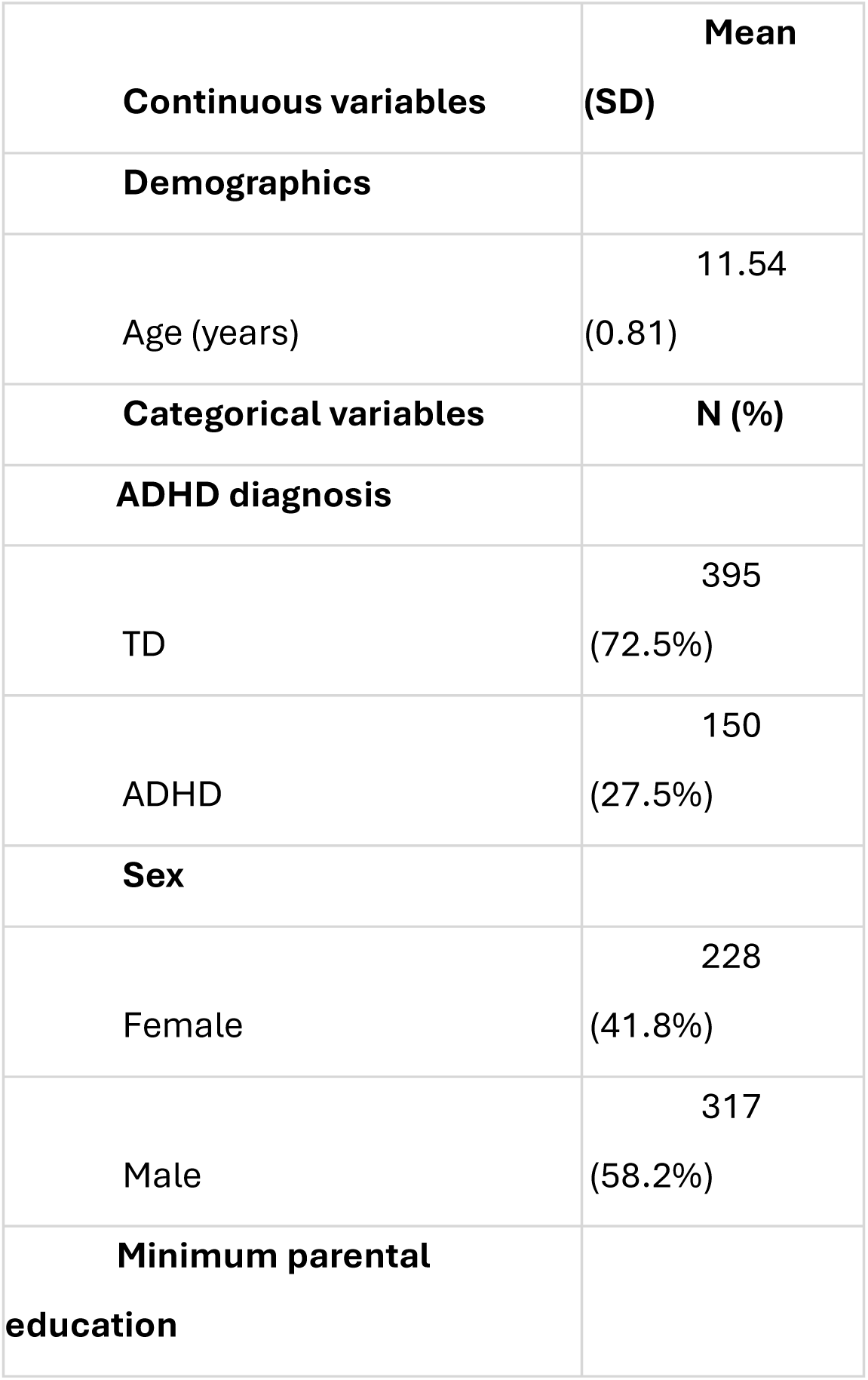
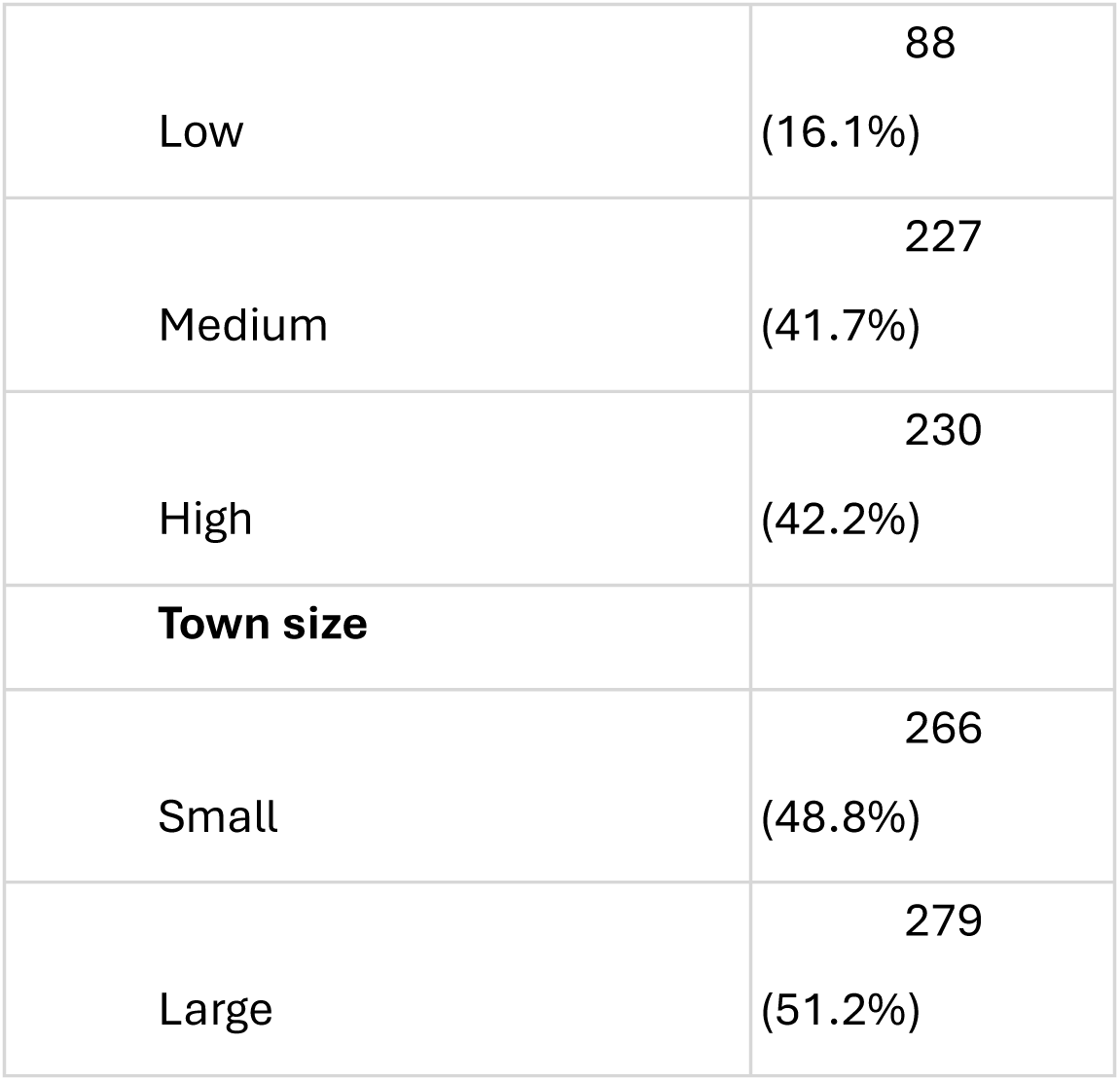
Descriptive statistics of the analytical sample. Abbreviations: ADHD, Attention Deficit/Hyperactivity Disorder; TD, typically developing; SD, standard deviation.

| **Continuous variables** | **Mean**<br>**(SD)** |
| --- | --- |
| **Demographics** |  |
| Age (years) | 11.54<br>(0.81) |
| **Categorical variables** | **N (%)** |
| **ADHD diagnosis** |  |
| TD | 395<br>(72.5%) |
| ADHD | 150<br>(27.5%) |
| **Sex** |  |
| Female | 228<br>(41.8%) |
| Male | 317<br>(58.2%) |
| **Minimum parental**<br>**education** |  |
| Low | 88<br>(16.1%) |
| Medium | 227<br>(41.7%) |
| High | 230<br>(42.2%) |
| **Town size** |  |
| Small | 266<br>(48.8%) |
| Large | 279<br>(51.2%) |

### Brain clusters with inhibition-related activation

Correct-rejection-versus-hit (i.e. correct NoGo vs. correct Go) contrast identified widespread and highly significant activation of the frontoparietal cognitive control network and the salience network (Table 3; Figure 3, upper panel). Significant clusters included bilateral activation in the Anterior Insula and a large medial cluster that encompassed the ACC and Pre-SMA. Additional activations were identified in the bilateral DLPFC and the bilateral Supramarginal Gyrus. Finally, a significant cluster was also observed in the Hippocampus.

**Figure 3.**
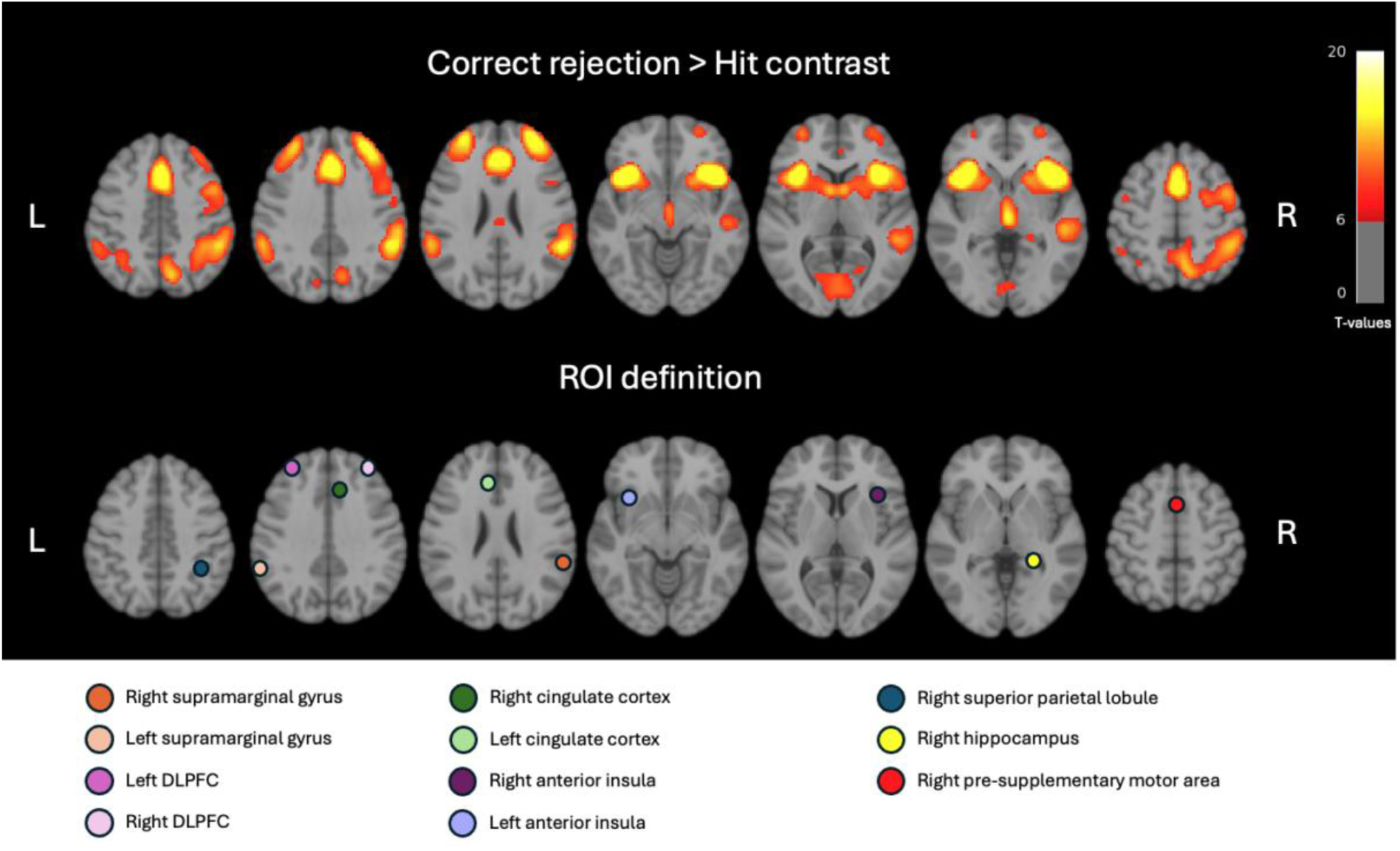
Maps of brain activity seen in the correct-rejection-versus-hit contrast from the Go/NoGo task (upper row). Voxels are color coded by their group-level t-values. The bottom row shows the placement of the 6 mm ROI spheres over whose voxels the average t-contrasts for the ROI analysis were computed. Abbreviation: DLPFC, dorsolateral prefrontal cortex. Threshold (upper panel): p<0.001 voxel-wise, p<10^-6^ FWE cluster-wise.

**Table 2.** Mean concentrations of air pollutants by time period in μg/m^3^. Abbreviations: PM_10_, Particulate matter with a diameter ≤ 10μm; NO_2_, Nitrogen dioxide; SD, standard deviation.

| Pollutant | Period | Mean (SD) |
| --- | --- | --- |
| PM<sub>10</sub> | Prenatal | 45.40<br>(14.43) |
|  | Early-life | 45.07 (6.59) |
|  | Current | 36.56 (4.90) |
| NO<sub>2</sub> | Prenatal | 23.84 (8.81) |
|  | Early-life | 24.48 (7.42) |
|  | Current | 19.18 (6.64) |

**Table 3.** Clusters showing inhibition-related activation (correct rejection/NoGo vs hit i.e. correct Go). For each cluster of voxels, the anatomical brain region, hemisphere, and number of voxels are listed, as well as the MNI coordinates and t-value of the cluster voxel with peak activation. Abbreviations: Hemisphere L/R, Left and Right; ACC, Anterior Cingulate Cortex; Pre-SMA, pre-supplementary motor area; DLPFC, dorsolateral prefrontal cortex; MNI, Montreal Neurological Institute.

| Cluster Region | Cluster Hemisphere | Cluster Size | Cluster voxel with peak activation |  |  |  |
| --- | --- | --- | --- | --- | --- | --- |
|  |  |  | x | y | z | t-value |
| Anterior Insula | L | 1137 | -36 | 18 | -8 | 20.69 |
| Anterior Insula | R | 1578 | 35 | 21 | 6 | 18.82 |
| ACC | R | 4288 | 9 | 25 | 35 | 15.86 |
| ACC | L | 4288 | -8 | 30 | 25 | 12.73 |
| Pre-SMA | R | 4288 | 2 | 11 | 52 | 13.22 |
| DLPFC | L | 753 | -32 | 45 | 35 | 11.78 |
| DLPFC | R | 1117 | 35 | 42 | 32 | 16.0 |
| Supramarginal Gyrus | L | 484 | -60 | -46 | 32 | 10.92 |
| Supramarginal Gyrus | R | 2272 | 59 | -41 | 28 | 15.18 |
| Superior Parietal Lobule | R | 2272 | 38 | -44 | 44 | 9.59 |
| Hippocampus | R | 52 | 23 | -37 | -1 | 9.79 |

### Exposure to air pollution and ROI activation

We found statistically significant negative relationships between current PM_10_ exposure and inhibition-related brain activity within several ROIs, specifically the bilateral DLPFC, bilateral ACC, bilateral anterior insula, bilateral superior parietal lobule, and bilateral supramarginal gyrus (Table 4 and Table S1). Furthermore, a significant negative effect of early-life PM_10_ exposure on brain activity was observed within the Pre-SMA.

**Figure 4.**
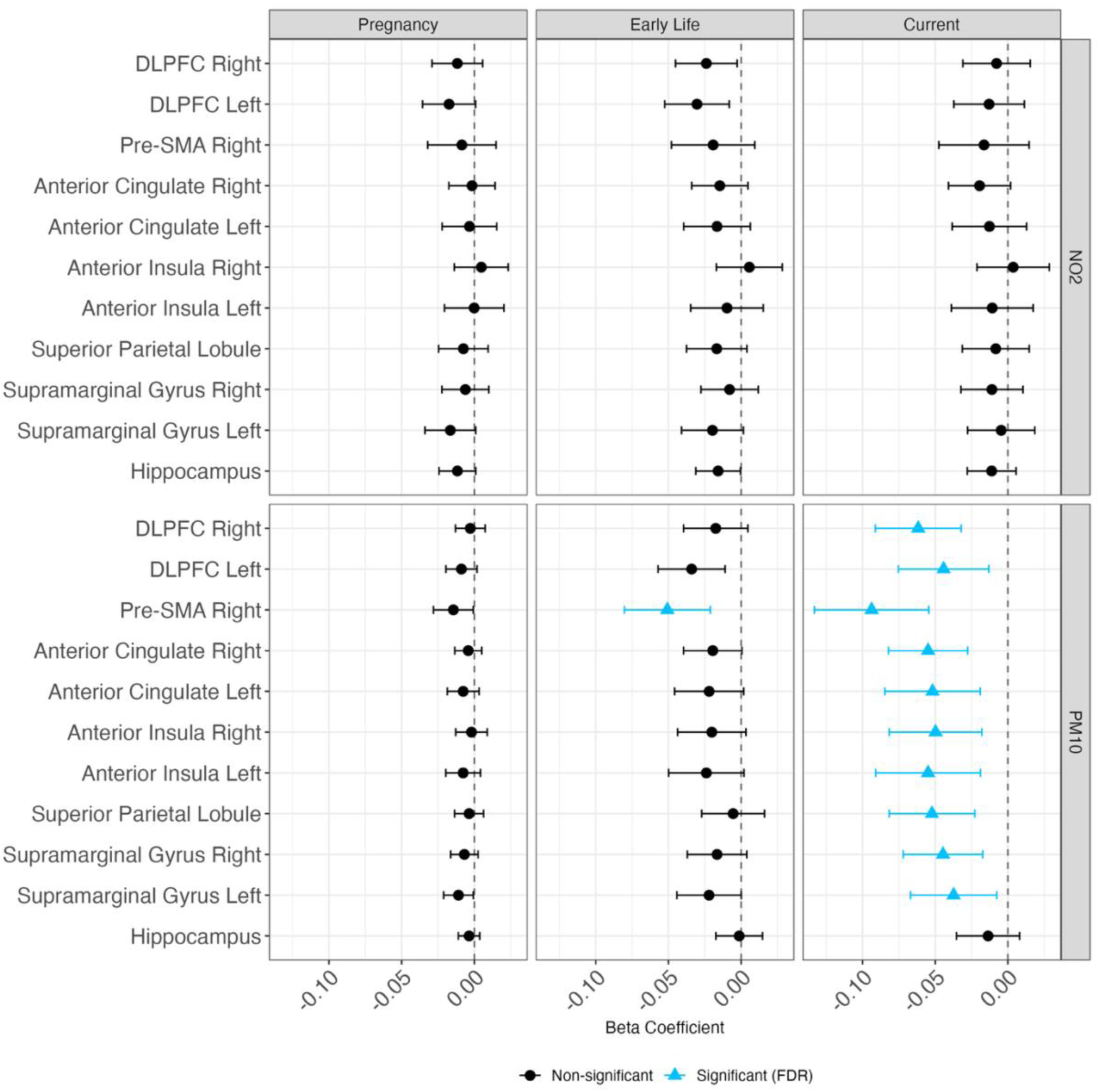
Beta estimates and 95% confidence intervals for the association between exposure to air pollution and inhibition-related brain activity for 11 regions of interest (ROIs).

To test the robustness these findings we ran several sensitivity analyses a) varying the size and the definition method for the ROIs and b) adding minimum parental education, a proxy for socioeconomic status to the models. All effects remained stable in models with ROI activations computed using alternative ROI sizes (Supplementary Figure S1 and S2). The only exception was the effect of early-life exposure to PM_10_ on the activity within the Pre-SMA, which lost significance when minimum parental education was included in the statistical model (Supplementary Figure S3) and when brain activity was computed over an ROI from all voxels from the entire anatomical region (Supplementary Figure S4.

Finally, none of the models/regions showed any significant interaction effect between ADHD status and exposure to air pollution (Supplementary Figure S5; all interaction p > 0.75).

## Discussion

Our study aimed to investigate the effects of long-term exposure to air pollution on brain activity during an inhibition fMRI task in 10-to 13-year-old children with and without ADHD. We found that higher current PM_10_ exposure was associated with decreased activity in key regions within the frontoparietal and salience networks—brain systems critical for executive functions and response inhibition (Aron et al., 2014; Botvinick et al., 2001; Cai et al., 2014; Corbetta & Shulman, 2002; Criaud et al., 2020; MacDonald, 2000; Mostofsky & Simmonds, 2008; Simmonds et al., 2008).

While other effects were mostly absent, we did observe a non-significant yet continuous pattern of decreased brain activity with higher early-life PM_10_ exposure, especially in the Pre-SMA, where the effect seemed to be the largest. The Pre-SMA is known to be involved in response selection and inhibition processes (Panek et al., 2025; Simmonds et al., 2008) and its decreased activation could explain the behavioral findings of less efficient conflict processing (Compa et al., 2023; Rivas et al., 2019), which may reflect impaired motor decisions (Panek et al., 2025). Concurrently, decreased activation within the salience and frontoparietal networks could lead to difficulties in task engagement and the ability to focus, respectively.

### Comparison with existing literature and future directions

To our knowledge, this is the first study linking exposure to air pollution and task-related brain activity. Most studies linking brain function and air pollution have focused on resting-state networks or structural abnormalities. While we did not perform connectivity analyses, we found decreased activity in similar attentional regions that other studies have reported as disrupted at rest (e.g., Cotter et al., 2024; Kusters et al., 2025). This consistency suggests that these networks are broadly vulnerable to air pollution’s effects.

Based on these findings and in accordance with studies regarding effects of exposure to air pollution on brain functional connectivity (Cotter et al., 2023; Gawryluk et al., 2023; Kusters et al., 2025), we hypothesize that connectivity within the frontoparietal and other attentional networks could also be decreased with higher exposures to air pollution. Consequently, a possible future direction in our data analysis will be to perform a network connectivity analysis within the frontoparietal and salience networks. Decreased connectivity within these task-positive networks could be related to less efficient information processing.

### Possible biological mechanisms

Our findings on decreased brain activity associated with exposure to air pollution can be explained by several plausible biological mechanisms through which air pollutants affect the brain. Pollutants can enter the brain either by crossing the blood-brain barrier via the bloodstream or through a direct pathway from the nasal passages to the olfactory bulb and prefrontal cortex (Gale et al., 2020; Manzano-Covarrubias et al., 2023; Wright & Ding, 2016). Once in the brain, they can trigger a cascade of neurotoxic events.

Particulate matter and other pollutants can activate microglia, the brain’s innate immune cells (R. Babadjouni et al., 2018; R. M. Babadjouni et al., 2017; Herting et al., 2024). Once activated, microglia become a source of pro-inflammatory cytokines and also generate reactive oxygen species (ROS) (Block & Calderón-Garcidueñas, 2009). This combined inflammatory–oxidative response can damage neurons and disrupt large-scale networks. microglia-driven inflammation and ROS production could contribute to the decrease in brain activity we found. Prolonged oxidative stress can impair cellular components and compromise neuronal integrity, potentially leading to functional reductions in affected brain areas.

In addition, particulate matter can weaken vascular barriers. Liu et al., (2021) demonstrated that exposure increases blood–brain barrier (BBB) permeability and vascular inflammatory signaling, particularly under reduced cerebral perfusion. Such BBB disruption facilitates the entry of circulating toxins and inflammatory molecules into the brain, amplifying neuroinflammatory and oxidative damage These biological mechanisms are supported by studies showing that exposure to air pollution is linked to structural damage to the brain, including reduced gray and white matter volume (Erickson et al., 2020; Huuskonen et al., 2021; Lubczyńska et al., 2020; Sukumaran et al., 2023; Szwed et al., 2025) and altered functional brain connectivity (Cotter et al., 2023; Gawryluk et al., 2023; Glaubitz et al., 2022; Pujol et al., 2016; Zundel et al., 2024) (see Morrel et al., 2025 for a systematic review of both). Our findings on reduced task-related brain activity align with this body of evidence, suggesting that these structural and functional changes may be the underlying cause of the less efficient information processing observed in our study.

### Air pollution, attention, the brain and ADHD

Multiple studies have reported links between exposure to air pollution and ADHD incidence in children (Kim et al., 2021; Markevych et al., 2018; Min & Min, 2017; Rosi et al., 2023; Thygesen et al., 2020). In the present study, we sought to examine whether ADHD diagnosis moderates the relationship between air pollution and brain activity during the Go/NoGo task and to test the hypothesis that children with ADHD, an at-risk population might be more vulnerable to detrimental effects of air pollution. We did not find any significant interaction terms for ADHD status and exposure to air pollution. On the contrary, consistent with another study from the same cohort (Lewandowska et al., 2025) we found that the brains of children with ADHD diagnosis and their typically developing peers were similarly affected. We therefore propose that the same mechanisms that underlie pollution’s effects on the brain and its attentional circuits may lead to the increased incidence of ADHD.

## Strengths & limitations

Our study has several strengths. Firstly, we used a randomly stratified sample of TD children, thus limiting selection bias. Secondly, all children’s MRI scans were acquired at one scanner site, which eliminated inter-scanner variability. Finally, we obtained complete lifelong addresses and used state-of-the-art air pollution modelling techniques to obtain maps of air pollution concentrations at a high spatial and temporal resolution.

This study also has some limitations. Firstly, MRI data was not available for all the children enrolled in the study, due to low quality of image acquisition and participants resigning during MRI scanning. Secondly, since air pollution modelling was only done for the study area, we lacked exposure data for children whose families had lived outside the study area during the prenatal or early-life periods. This was also the reason why the sample size for analyses of current air pollution exposure was slightly larger.

## Conclusions

Our study found that higher long-term exposure to PM_10_ is associated with decreased brain activity in regions involved in key attentional and executive networks. These findings add novel, pertinent evidence to the growing body of research showing that air pollution can impair executive function development in children (Compa et al., 2023; Forns et al., 2016, 2017; Gignac et al., 2021, 2022; Guxens et al., 2014; Saenen et al., 2016; Sunyer et al., 2015, 2017). We propose that these impairments may arise from disrupted information processing within the frontoparietal and salience networks, which are critical for executive and attentional control. Moving beyond existing structural and resting-state studies, our work provides critical insights into how air pollution affects executive processes in children.

## CRediT authorship contribution statement

**Mikołaj Compa:** Conceptualization, Methodology, Software, Formal analysis, Data curation, Writing – original draft, Writing – review & editing, Visualization

**Yarema Mysak:** Data curation

**Aleksandra Domagalik-Pittner:** Writing – review & editing, Investigation, Resources

**Bartosz Kossowski:** Software

**Paulina Lewandowska:** Writing – review & editing

**Clemens Baumbach**: Data curation, Writing – Review & Editing

**Katarzyna Kaczmarek-Majer**: Software, Resources

**Krzysztof Skotak**: Resources

**Anna Degórska**: Software, Resources

**Katarzyna Sitnik-Warchulska**: Investigation, Resources

**Małgorzata Lipowska**: Investigation, Resources

**Bernadetta Izydorczyk**: Investigation, Resources

**James Grellier**: Writing – review & editing

**Iana Markevych**: Resources, Data curation, Writing – review & editing, Supervision

**Marcin Szwed**: Conceptualization, Methodology, Resources, Writing – review & editing, Funding acquisition, Supervision

## Data Availability

All data produced in the present study are available upon reasonable request to the authors

## Acknowledgments

We are very grateful to all of the children and their parents for their participation in the study. We thank our partner schools for their help in recruiting the typically developing group of children. We also extend our gratitude to the field psychologists, who identified children with ADHD and performed psychological testing on all children. The efforts of the study team in providing technical, logistic, administrative, and communication support were invaluable. We also thank the group of our Master’s students who helped in the acquisition of MRI data, as well as our MRI technicians for their hard work.

